# Predictors of COVID-19 Vaccine Hesitancy and Adverse Events following COVID-19 vaccination among People on Anti-retroviral therapy in Nigeria

**DOI:** 10.64898/2025.12.05.25341692

**Authors:** Victoria Etuk, Oluwafemi Omonijo, Charity Sanni, Stella Atema, Abimiku Alash’le, Evaezi Okpokoro

**Affiliations:** International Research Centre of Excellence, Institute of Human Virology, Nigeria; Institute of Human Virology, School of Medicine, University of Maryland, Baltimore

## Abstract

**Background:** COVID-19 vaccination is critical among people living with HIV (PLHIV) given the risk of severe COVID-19 disease and hospitalization, due to their immunocompromised state. However, COVID-19 vaccine hesitancy among PLHIV globally has been increasing, thus presenting a risk of continued transmission of the disease. We sought to investigate the rate of, and predictors of COVID-19 vaccine hesitancy among people living with HIV in FCT, Nigeria. Additionally, we sought to investigate adverse events following immunization among PLHIV who were already vaccinated.

**Methods:** A facility-based cross-sectional study was conducted among 164 unvaccinated PLHIV across two-large ART clinics in 2023. An interviewer administered data collection tool was utilized to collect data on socio-demographic information, vaccine hesitancy and barriers to vaccine uptake. Among the unvaccinated individuals, we adapted the WHO-SAGE definition to measure vaccine hesitancy, by asking “*Do you still want to take the vaccine*?”. Descriptive statistics, chi-squares and binary logistic regressions were used to analyse the data in Stata version 18.

**Results:** Majority of the unvaccinated individuals were female (79.3%), and aged 35-49 years (62.8%). Over half (62.2%, 104) were hesitant to receive a COVID-19 vaccine. On multi-variable logistic regression, the odds of vaccine hesitancy was 5 times higher among participants without pre-existing co-morbidities (AOR: 4.858, 95%CI: 1.109,21.285, p=0.036). Barriers to vaccination among unvaccinated, non-hesitant participants included not knowing available vaccination centers (40.7%, 24/59), and lack of transportation to a vaccination center (22%, 13/59). Among participants who were vaccinated, 45 (52.9%) reported at least one adverse event (AE). Most common AEs reported were local reactions (15, 32.6%), muscle pain (13, 28.2%), fever (10, 21.7%) and malaise (7, 15.2%). The odds of AE were higher among females (OR=3.978, p=0.004). Age, viral load, antiretroviral regimen and presence of other comorbidities did not affect AE occurrence (p-value for all >0.05). No serious AE was reported.

**Conclusion:** COVID-19 Vaccine hesitancy rate was high among our study population. Measures to address vaccine hesitancy among PLHIV should be prioritized by immunization and HIV programs.

## Introduction

Vaccinations are the most potent means of prevention of COVID-19 morbidity and mortality[1–3], despite the promotion of non-pharmaceutical interventions at the onset of the pandemic. [4], Significant efforts have been made towards accelerated development of vaccines[5], however only, 12 vaccines have been approved for emergency licensure by WHO[6], with vaccine effectiveness ranging from 62% to 98%, across different vaccine types (mRNA, viral vector and adenoviruses), and doses. [7,8].

COVID-19 vaccination is critical among people living with HIV (PLHIV) as they have an increased risk of severe COVID-19 disease and hospitalization[9], due to their immunocompromised state. However, globally COVID-19 vaccine acceptance in this population has been low, with vaccine acceptance reported to be about 62-67% [10,11] and uptake reported at about 56.6%[11], despite proven safety and effectiveness of the vaccines among PLHIV[12–14]. In Nigeria, COVID-19 vaccine acceptance among PLHIV is also reportedly low, at about 46%, as compared with the target uptake of 70%[15,16]. These low acceptance and uptake rates have been strongly associated with vaccine hesitancy among PLHIV.

Vaccine hesitancy is defined as the refers to “delay in acceptance or refusal of vaccination despite availability of vaccination services”[17]. There has been a rise in COVID-19 vaccine hesitancy globally, which has been associated with an increase in COVID-19 associated infodemics[18], and fear of adverse events [19], thus presenting a risk of continued transmission of the disease, in the midst of newer variants of the virus. Our study sought to investigate the prevalence and predictors of COVID-19 vaccine hesitancy and adverse events following vaccination among people living with HIV, and barriers to COVID-19 vaccination in this population.

## Methods

### Study design, setting, and participants

We have previously reported on COVID-19 vaccine uptake in this population. This study was a multi-centre cross-sectional study conducted in the FCT[20]. The study was conducted in two large clinics providing antiretroviral care- the Asokoro District Hospital and the Kubwa General Hospital, from April to July, 2023 (Figure 1).

**Figure 1:**
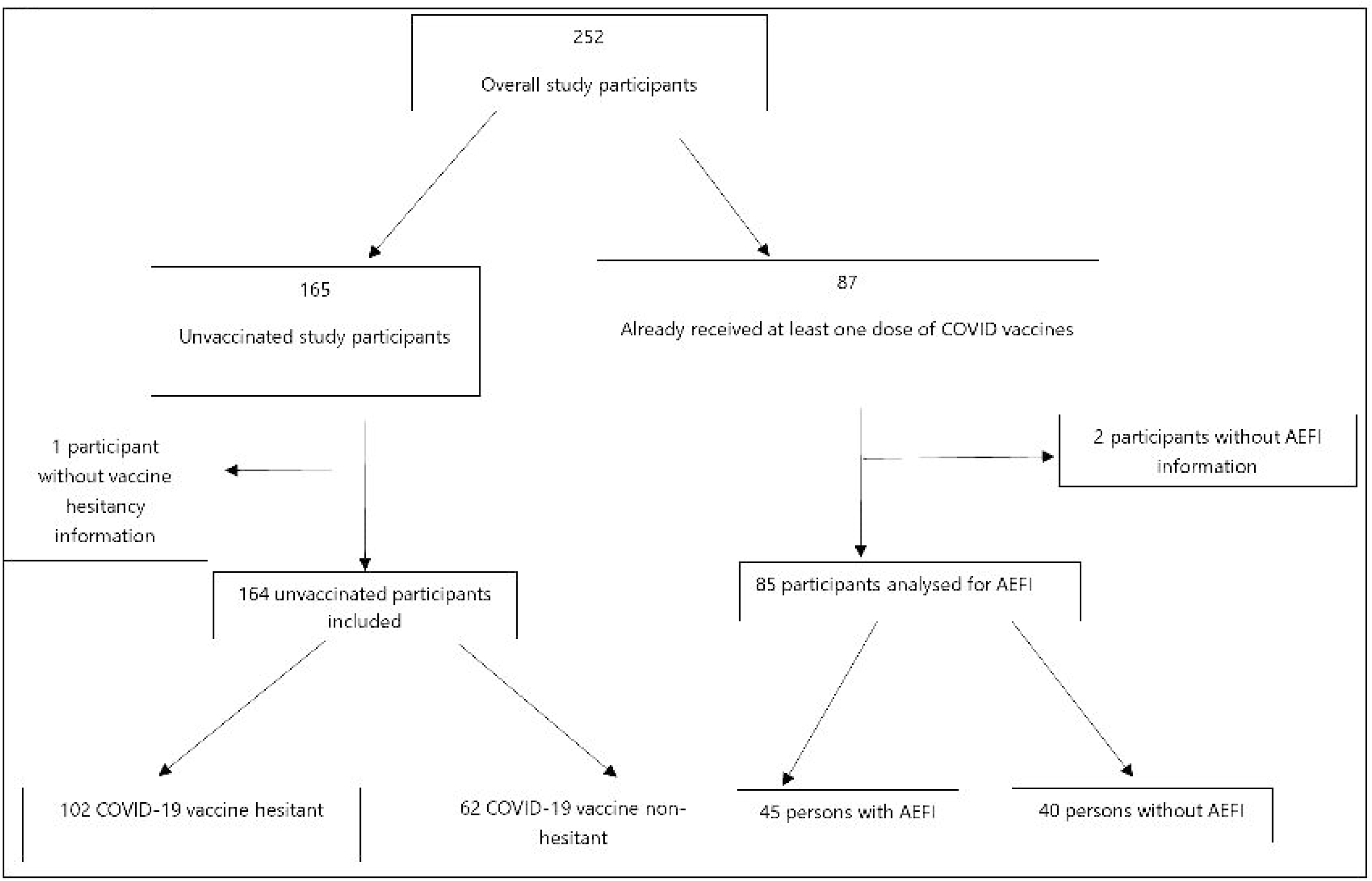
Study Schema.

### Inclusion and Exclusion criteria

Participants who were included in the analysis were those with confirmed HIV status and were at least 18 years old and on ART for at least 6 months. Participants who were participating in other studies administering COVID-19 vaccines were excluded (Figure 1).

### Sample size calculations and sampling

We estimated a sample size of 252 participants based on the Leslie-Kish formula for cross-sectional studies, of which 164 participants (65.1%) had not received any dose of COVID-19 vaccine and included in the analysis for vaccine hesitancy, while only vaccinated individuals were analysed for AEFI information. Thus, the outcome variable for vaccine hesitancy was only measured among the 164 participants who had not received any COVID-19 vaccines (Figure 1).

### Data Collection

We administered a 50-item, interviewer-administered questionnaire to participants. The questionnaire included questions on socio-demographic information, COVID-19 vaccine uptake, barriers to COVID-19 vaccination and AEFI experience. Additional clinical information on ART characteristics (ART regimen, duration of ART regimen, CD4 count and viral load) were obtained from participants hospital case notes and electronic medical records.

### Variables

#### Outcome variables

##### COVID-19 vaccine hesitancy

The main outcome variable was COVID-19 vaccine hesitancy. Participants were who reported not having received any dose of a COVID-19 vaccine were asked the question “*Do you still want to take the vaccine?”.* Those who responded ‘No’ were deemed to have refused the COVID-19 vaccine and were considered COVID-19 vaccine hesitant.

##### Barriers to COVID-19 vaccine uptake

Non-hesitant participants who responded “yes” to “*Do you still want to take the vaccine?”*, were further asked *‘What makes it difficult for you to get a COVID-19 vaccine’?*

##### Adverse events following COVID-19 Vaccine

Among participants who had received vaccines, information on side effects experienced were also collected.

#### Independent variables

The independent variables included socio-demographic information (age, sex, level of education, income e.t.c), ART information (ART regimen, duration of ART regimen, CD4 count and viral load) and clinical characteristics (previous vaccine history and presence of co-morbidities).

### Ethical Approvals

The Federal Capital Territory Health Research Ethics Committee, and the research ethics committees of both study sites approved the study (Approval Number FHREC/2023/01/35/15-03-23; REC/KubwaGH/2023/04/17/01; FCTA/HHSS/HMB/ADH/133/23). We obtained written informed consent from all participants.

### Data Management and Analysis

Data was collected and stored on REDCap database and analysed using Stata version 18. Descriptive analyses were conducted for outcome and independent variables using frequencies and proportions. Chi-square tests were used to conduct bivariate analysis of the outcome and independent variables. Variables with p-values less than 0.2 on the bivariate analysis were included in a multivariate, binary logistic regression. Independent variables with p values <0.05 in the logistic regression model were adjudged to be statistically significant and presented with odds ratios and 95% confidence intervals.

## Results

Table 1 shows the socio-demographic and clinical characteristics of the 164 study participants. Majority of the study participants were female (79.3%) and aged 35-49 years (62.8%). About 10% of the females were pregnant, and over three-quarters (75.6%) had at least secondary education and were employed (78.0%). Majority were Christian (81.6%) and resided in semi-urban settings (68.9%).

**Table 1:** Baseline Characteristics of Study Participants.

|  | **Baseline**<br>**Characteristic** | **Frequency**<br>**N=164** | **Percentage (%)** |
| --- | --- | --- | --- |
| **Socio-demographic** |  |  |  |
| Age group | 18-34 | 38 | 23.2 |
|  | 35-49 | 103 | 62.8 |
|  | 50 and above | 23 | 14.0 |
| Sex | Female | 130 | 79.3 |
|  | Male | 34 | 20.7 |
| Pregnancy status (female participants) | Pregnant | 13 | 10.0 |
|  | Not pregnant | 117 | 90.0 |
| Educational status | Less than secondary | 40 | 24.4 |
|  | Secondary and above | 124 | 75.6 |
| Marital Status | Single | 35 | 21.3 |
|  | Married | 85 | 51.8 |
|  | Divorced | 4 | 2.4 |
|  | Widowed | 31 | 18.9 |
|  | Separated | 9 | 5.5 |
| Monthly Income (naira) | Less than 10,000 | 41 | 25.0 |
|  | 10,000-50,000 | 73 | 44.5 |
|  | 50,000- 100,000 | 35 | 17.6 |
|  | Greater than 100,000 | 15 | 9.1 |
| Residence | Rural | 51 | 31.1 |
|  | Semi-Urban | 109 | 68.9 |
| Employment status | Unemployed | 36 | 22.0 |
|  | Employed | 128 | 78.0 |
| Religion | Christianity | 133 | 81.6 |
|  | Islam | 30 | 18.4 |
| Facility | Asokoro District Hospital | 78 | 47.6 |
|  | Kubwa General Hospital | 86 | 52.4 |
| Number of persons living in household | 1 | 13 | 7.9 |
|  | 2-5 | 58 | 35.4 |
|  | Greater than 5 | 93 | 56.7 |
|  | Yes | 132 | 80.5 |
| **ART characteristics** |  |  |  |
| Duration on ART (years) | Less than 10 years | 132 | 80.5 |
|  | 10 years and above | 32 | 19.5 |
| Viral suppression | Virally suppressed | 138 | 85.7 |
|  | Virally unsuppressed | 23 | 14.3 |
| **Clinical characteristics** |  |  |  |
| Received other vaccines in adulthood | Yes | 68 | 41.5 |
|  | No | 96 | 58.5 |
| Type of vaccine received in adulthood | Yellow fever | 9 | 5.5 |
|  | Hepatitis B | 25 | 15.2 |
|  | Tetanus | 45 | 27.3 |
|  | Others | 4 | 2.4 |
| Presence of co-morbidities | Yes | 10 | 6.1 |
|  | No | 154 | 93.9 |
| Type of co-morbidity | Hypertension | 7 | 70.0 |
|  | Diabetes | 1 | 10.0 |
|  | Tuberculosis | 1 | 10.0 |
|  | Ulcer | 1 | 10.0 |

All participants were on anti-retroviral therapy, with most on ART for less than 10 years. Most participants were virally suppressed (85.7%). About half had received other adulthood vaccines (58.5%), while less than 10% of participants had other co-morbidities (Table 1).

### Vaccine hesitancy and associated factors

Overall, of the 164 unvaccinated participants, 62.2% (104) were hesitant to receive COVID-19 vaccines. Vaccine hesitancy was higher in PLHIV who had greater than secondary education (65.3%), were employed (65.6%), and accessed care at Asokoro District Hospital (70.5%). Similarly, female participants who were not pregnant had higher rates of COVID-19 vaccine hesitancy (64.1%). Vaccine hesitancy was higher among those who had received other vaccines in adulthood (73.5%). In contrast, COVID-19 vaccine hesitancy was lower among PLHIV who were on ART for less than 10 years (60.6%) and virally unsuppressed (56.5%). Facility of care (p=0.036), religion (0.046) and receiving other vaccines in adulthood (p=0.012) had statistically significant associations with vaccine hesitancy (Table 2).

**Table 2:** Factors Associated with vaccine hesitancy.

| Independent Variables |  | Hesitant<br>102<br>(62.2%) | Not<br>hesitant<br>62 (37.8%) | X2 | p-value |
| --- | --- | --- | --- | --- | --- |
| **Socio-demographic** |  |  |  |  |  |
| Age group | 18-34 | 24 (63.2) | 14 (36.8) | 0.0323 | 0.984 |
|  | 35-49 | 64 (62.1) | 39 (37.9) |  |  |
|  | 50 and above | 14 (60.9) | 9 (39.1) |  |  |
| Sex | Female | 81 (62.3) | 49 (37.7) | 0.0034 | 0.954 |
|  | Male | 21 (61.8) | 13 (38.2) |  |  |
| Educational status | Less than secondary | 21 (52.5) | 19 (47.5) | 2.1149 | 0.146 |
|  | Secondary and above | 81 (65.3) | 43 (34.7) |  |  |
| Marital Status | Single | 24 (68.6) | 11 (31.4) | 3.7945 | 0.443* |
|  | Married | 55 (64.7) | 30 (35.3) |  |  |
|  | Divorced | 3 (75.0) | 1 (25.0) |  |  |
|  | Widowed | 16 (51.6) | 15 (48.4) |  |  |
|  | Separated | 4 (44.4) | 5 (55.6) |  |  |
| Income | Less than 10,000 | 24 (58.5) | 17 (41.5) | 4.3300 | 0.228 |
|  | 10,000-50,000 | 41 (56.2) | 32 (43.8) |  |  |
|  | 50,000- 100,000 | 26 (74.3) | 9 (25.7) |  |  |
|  | Greater than 100,000 | 11 (73.3) | 4 (26.7) |  |  |
| Employment status | Unemployed | 18 (50.0) | 18 (50.0) | 2.9175 | 0.088 |
|  | Employed | 84 (65.6) | 44 (34.4) |  |  |
| Residence | Rural | 33 (64.7) | 18 (35.3) |  |  |
|  | Semi-Urban/Urban | 69 (61.1) | 44 (38.9) |  |  |
|  |  |  |  | 0.198 | 0.656 |
| Religion | Christianity | 88 (66.2) | 45 (33.8) | 3.9741 | 0.046 |
|  | Islam | 14 (46.7) | 16 (53.3) |  |  |
| Facility | Asokoro District Hospital | 55 (70.5) | 23 (29.5) | 4.3767 | 0.036 |
|  | Kubwa General Hospital | 47 (54.6) | 39 (45.5) |  |  |
| Number of persons living in household | 1 | 7 (53.9) | 6 (46.1) | 1.8853 | 0.390 |
|  | 2-5 | 40 (69.0) | 18 (31.0) |  |  |
|  | Greater than 5 | 55 (59.1) | 38 (40.9) |  |  |
| Pregnancy status (female participants) | Pregnant | 6 (46.2) | 7 (53.8) | 1.6049 | 0.205 |
|  | Not pregnant | 75 (64.1) | 42 (35.9) |  |  |
| ART characteristics |  |  |  |  |  |
| Duration on ART | Less than 10 years | 80 (60.6) | 52 (39.4) | 0.7265 | 0.394 |
|  | 10 years and above | 22 (68.8) | 10 (31.3) |  |  |
| Viral suppression | Virally suppressed | 86 (62.3) | 52 (37.7) | 0.2798 | 0.597 |
|  | Virally unsuppressed | 13 (56.5) | 10 (43.5) |  |  |
| Clinical characteristics |  |  |  |  |  |
| Received other vaccines in adulthood | Yes | 50 (73.5) | 18 (26.5) | 6.3470 | 0.012 |
|  | No | 52 (54.2) | 44 (45.8) |  |  |
| Presence of co-morbidities | Yes | 4 (40.0) | 6 (60.0) | 2.2312 | 0.135* |
|  | No | 98 (63.6) | 56 (36.4) |  |  |

\*Fisher's exact

On multivariate logistic regression, the most significant predictor of COVID-19 vaccine hesitancy was history of co-morbidities. The odds of vaccine hesitancy was 5 times more among participants who had no-comorbidities as compared to those with co-morbidities [AOR=4.858, 95%CI= 1.109,21.285, p-value= 0.036] (Table 3). Participants with at least secondary school education [AOR=0.906], were Muslim [AOR=0.448], accessed care at Kubwa General Hospital [AOR=0.679] and had not taken other vaccines in adulthood [AOR=0.582] had lower odds of vaccine hesitancy, although the associations were not statistically significant. Conversely, participants who were employed [AOR=1.208] and not pregnant [AOR=1.878] had higher odds of vaccine hesitancy, but these associations were not statistically significant (Table 3).

**Table 3:** Logistic regression factors associated with COVID-19 hesitancy.

| Independent Variable |  | COR | 95%CI | p-value | AOR | 95% CI | p-value |
| --- | --- | --- | --- | --- | --- | --- | --- |
| Educational status | Less than secondary (Ref) |  |  |  |  |  |  |
|  | Secondary and above | 1.704 | 0.828,3.510 | 0.148 | 0.918 | 0.357,2.359 | 0.860 |
| Employment status | Unemployed (Ref) |  |  |  |  |  |  |
|  | Employed | 1.909 | 0.903,4.034 | 0.090 | 1.231 | 0.489,3.106 | 0.658 |
| Pregnancy status (female participants) | Pregnant (Ref) |  |  |  |  |  |  |
|  | Not pregnant | 2.083 | 0.657,6.606 | 0.213 | 1.789 | 0.521,6.139 | 0.355 |
| Religion | Christianity (Ref) |  |  |  |  |  |  |
|  | Islam | 0.447 | 0.201,0.998 | **0.049** | 0.495 | 0.149,1.642 | 0.251 |
| Facility | Asokoro District Hospital (Ref) |  |  |  |  |  |  |
|  | Kubwa General Hospital | 0.504 | 0.264,0.961 | **0.038** | 0.642 | 0.278,1.476 | 0.297 |
| Presence of co-morbidities | Yes (Ref) |  |  |  |  |  |  |
|  | No | 2.625 | 0.710, 9.70 | 0.148 | 4.858 | 1.109,21.285 | **0.036** |
| Received other vaccines in adulthood | Yes (Ref) |  |  |  |  |  |  |
|  | No | 0.425 | 0.217,0.833 | **0.013** | 0.591 | 0.263,1.324 | 0.201 |

*\*Values in bold are significant at 5% level of significance*

### Barriers to COVID-19 vaccine among non-hesitant participants

Figure 2 shows the barriers to COVID-19 vaccination among non-hesitant participants. Majority of the participants (40.7%, 24/59) did not know where to get vaccinated. Others reported lack of transportation to a vaccination centre and reported that they had physical limitations (22.0% vs 10.2% respectively).

**Figure 2:**
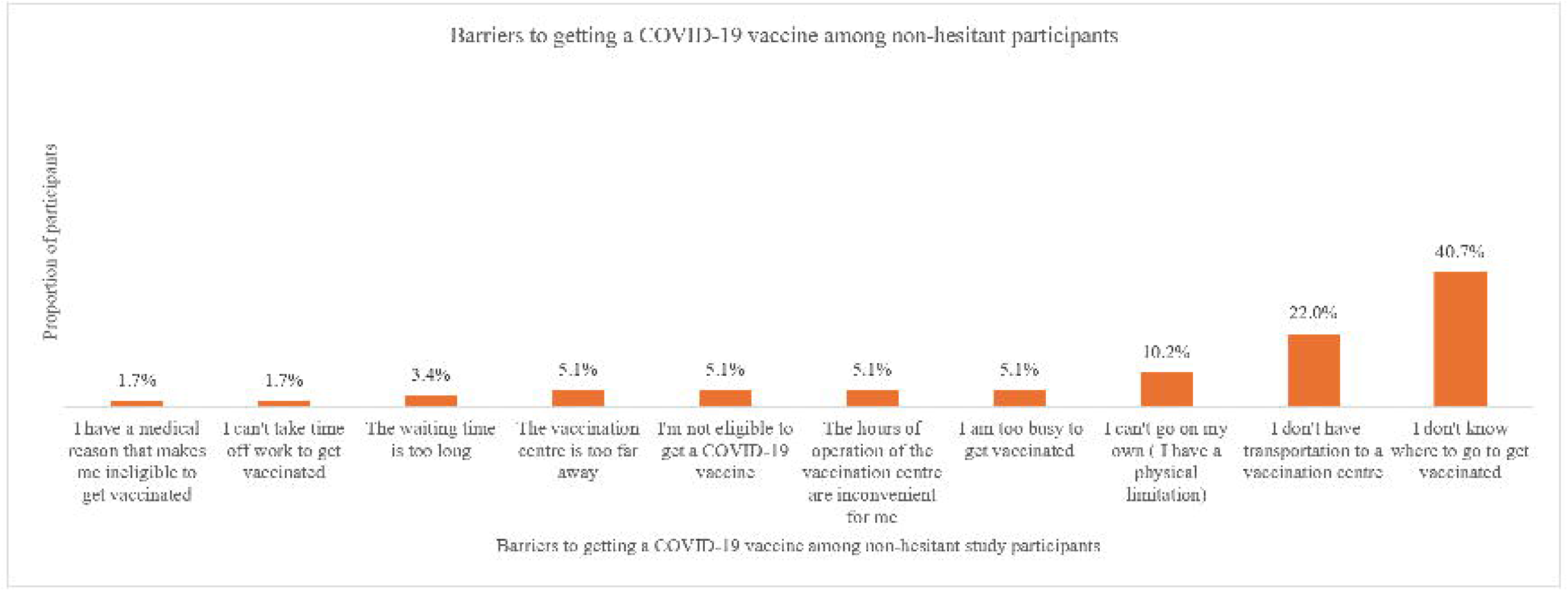
Barriers to COVID-19 vaccination

### Adverse Events following COVID-19 vaccination among vaccinated participants

Of the 87 vaccinated participants, 46 (52.8%) reported at least one AE. Most common AEs reported were local reactions (15, 32.6%), muscle pain (13, 28.2%), fever (10, 21.7%) and malaise (7, 15.2%) (Figure 3). The odds of AEs were higher among females (OR=3.978, p=0.004) (Table 4). Age, viral load, antiretroviral regimen and presence of other co-morbidities did not affect AE occurrence (p-value for all >0.05) (Table 4). No serious AE was reported.

**Figure 3:**
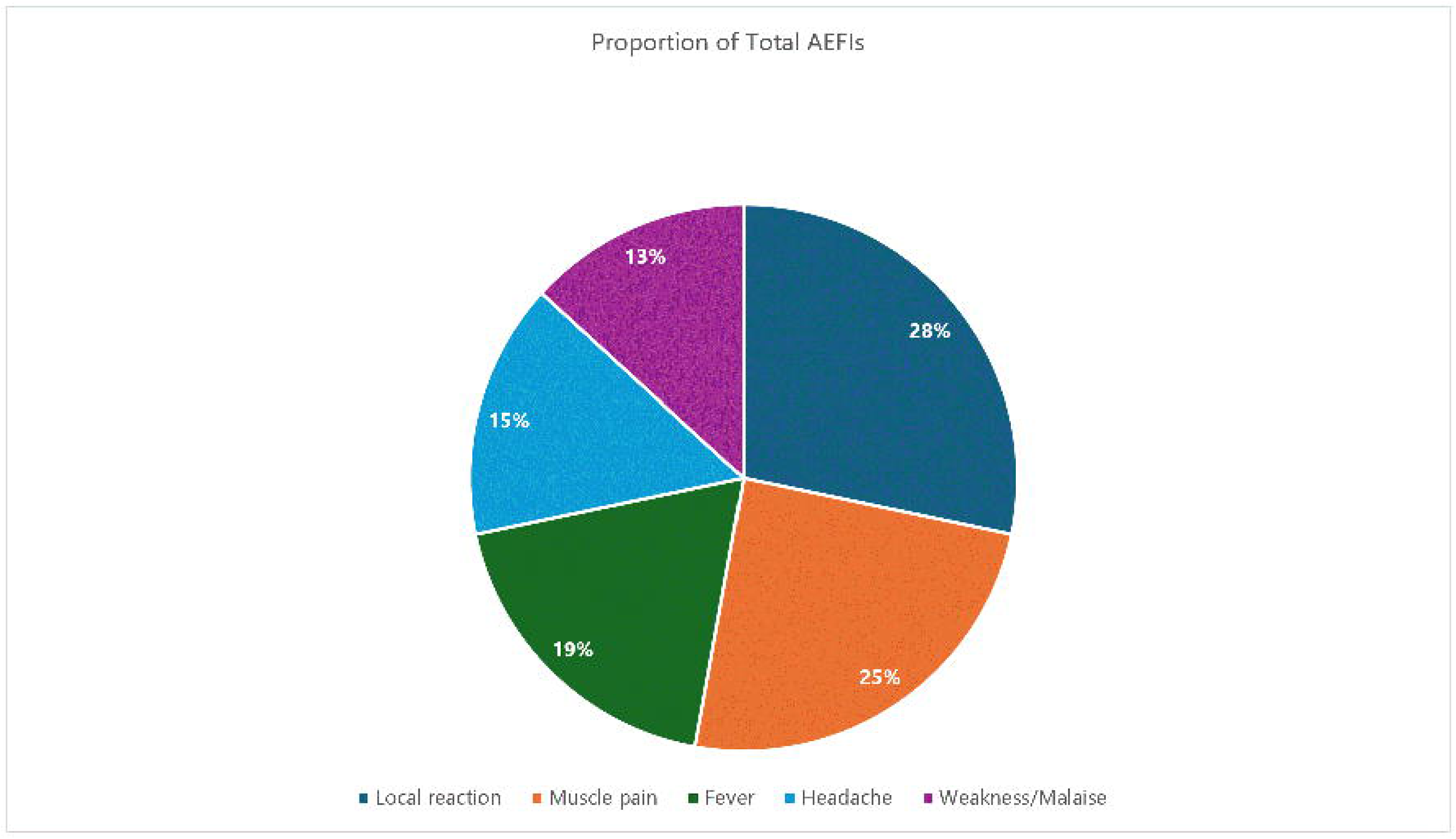
Type of AEFIs reported (N=45)

**Table 4:** Predictors of AEFI among PLHIV in FCT, Nigeria.

| Characteristic | Total | AEFI reported | No AEFI reported | COR | p-value |
| --- | --- | --- | --- | --- | --- |
| **Age** |  |  |  |  |  |
| **18-34** | 8 | 2(75.0) | 6 (25.0) | Ref | 0.123 |
| **35-49** | 52 | 29(55.8) | 23 (44.2) | 3.782 (0.697, 20.526) | 0.141 |
| **50 and above** | 25 | 14 (56.0) | 11 (44.0) | 3.818 (0.141,22.744) |  |
| **Sex** |  |  |  |  |  |
| **Female** |  | 36 (65.5) | 19 (34.5) | 4.42 (1.695, 11.529) | 0.002 |
| **Male** | 55 | 9 (30.0) | 21 (70.0) | Ref |  |
|  | 30 |  |  |  |  |
| **Duration on ART** |  |  |  |  |  |
| **Less than 10 years** | 58 | 31 (53.5) | 27 (46.5) | 1.148 (0.454, 2.897) |  |
| **10 years and above** | 26 | 13 (50.0) | 13 (50.0) | Ref | 0.770 |
| **ART regimen** |  |  |  |  |  |
| **DTG-based** |  | 44 (53.0) | 39 (47.0) |  |  |
| **Non DTG-based** | 83 | 1 (50.0) | 1 (50.0) |  |  |
|  | 2 |  |  |  |  |
| **Viral suppression** |  | 40 (50.6) | 30 (49.4) | 0.342 (0.034, 3.430) |  |
| **Virally suppressed** | 79 |  |  |  |  |
| **Virally unsuppressed** | 4 | 3 (75.0) | 1 (25.0) | Ref |  |
| **Co-morbidities** |  |  |  |  |  |
| **Yes** | 8 | 5 (62.5) | 3 (37.5) | 1.541 (0.344, 6.906) | 0.572 |
| **No** | 77 | 40 (51.9) | 37 (48.1) | Ref |  |
| **Vaccination status** |  |  |  |  |  |
| **Fully vaccinated** | 57 | 31 (54.4) | 26 (45.6) | 0.838 (0.339, 2.074) | 0.704 |
| **Partially vaccinated** | 28 | 14 (50.0) | 14 (50.0) | Ref |  |

## Discussion

Our study sought to investigate COVID-19 vaccine hesitancy and its associated factors among PLHIV. There was a high rate of COVID-19 vaccine hesitancy in our population (62.2%), similar to other reported studies in Nigeria (57.7%)[21], however it was higher than those reported in other African countries (21%-27%), and other high income countries [10,22,23]. Vaccine hesitancy rates in this population was also similar to those reported in general populations [24,25]. High vaccine hesitancy rates in this study is concerning and may be an indication that COVID-19 vaccination campaigns and awareness programmes did not specifically target PLHIV. Similarly, it may be an indicator that vaccination campaigns were not effective in improving COVID-19 vaccine acceptance among PLHIV.

Vaccine hesitancy rates in our study differed by religion, facility and previous vaccine history, with Christians being more vaccine hesitant than muslims. It is well established that religious beliefs affects vaccine hesitancy, with COVID-19 vaccine in particular [26–28]. It is therefore critical to involve religious leaders in combating vaccine hesitancy among both the general population and PLHIV. Convincing religious leaders of the safety and effectiveness of the COVID-19 vaccine and involving them in vaccine uptake messaging, will reduce vaccine hesitancy, particularly among PLHIV.

The presence of co-morbidities was the most significant predictor of COVID-19 vaccine hesitancy in our study. Participants who had no co-morbidities were 5 times more likely to be vaccine hesitant, compared to those without co-morbidities. It has been well established that the risk of COVID-19 disease is higher in the presence of pre-existing co-morbidities like hypertension, diabetes and obesity, both in people with and without HIV [29], thus people without these co-morbidities may be vaccine hesitant as they may have a lower risk perception of COVID-19 disease.

Previous vaccine history was associated with vaccine hesitancy in our setting. There was however a higher rate of vaccine hesitancy among previously vaccinated PLHIV, in contrast to other studies [30]. A possible explanation may be the rise of COVID-19 associated infodemics, which may propel vaccine hesitancy even among previously vaccinated participants.

We observed that a high proportion of non-hesitant PLHIV in our study did not know where to get vaccinated, despite COVID-19 vaccines being available at no cost in health facilities across Nigeria [31]. This demonstrates a need for the expanded programme on immunization at national and sub-national levels to have COVID-19 vaccine awareness campaigns targeted for PLHIV, with an emphasis on vaccination sites. A possible solution may be to have vaccination outlets at ART clinics to drive awareness and uptake. These results also beyond creating awareness, immunization programmes should also incorporate messages on vaccination sites and availability knowledge of a vaccine during vaccination campaigns.

Although a high proportion of vaccinated individuals reported AEFIs, all AEFIs were mild and no serious AEFI was reported. This is similar to reports of COVID-19 associated AEFIs among both PLHIV (32) and the general population(32). This demonstrates the safety of COVID-19 vaccines in this population, however, this study relied on self-reports of AEFI, and study participants who reported AEFI may have recall bias, as the AEFI information was provided retrospectively.

## Limitations

The study did not account for reasons for vaccine hesitancy and did not measure possible changes in hesitancy through repeated measures. Further studies can measure changes in vaccine hesitancy over time among PLHIV and explore reasons for vaccine hesitancy. Additionally, the AEFI data was collected retrospectively, and may have been underreported due to recall bias.

## Conclusion

Vaccine hesitancy was high in our study population and was associated with presence of co-morbidities. Targeted messaging and vaccine campaigns can help reduce vaccine hesitancy among PLHIV.

## Data Availability

All data produced in the present study are available upon reasonable request to the authors

## Authors’ contributions

Victoria Peter Etuk and Evaezi Okpokoro conceived the idea and designed the study; Victoria Peter Etuk, Oluwafemi Omonijo, Charity Sanni and Stella Atema conducted the field implementation and data collection. Victoria Peter Etuk carried out the data analyses and visualisations. Victoria Peter Etuk wrote the initial draft of the manuscript. Victoria Peter Etuk, Oluwafemi Omonijo, Charity Sanni, Stella Atema, Evaezi Okpokoro and Alash’le Abimiku PhD carried out the critical review of the manuscript. All the authors have read and approved the final version of this manuscript.

## Acknowledgements

The authors are grateful to the management of the Asokoro District Hospital and Kubwa General Hospital for their support for the study. The efforts of Christie Omali and Chiamaka Akuma-Akpu as research assistants on the study are appreciated.

